# From classic to rap: Airborne transmission of different singing styles, with respect to risk assessment of a SARS-CoV-2 infection

**DOI:** 10.1101/2021.03.25.21253694

**Authors:** Bernhard Richter, Anna Hipp, Bernd Schubert, Marcus Rudolf Axt, Markus Stratmann, Christian Schmölder, Claudia Spahn

**Affiliations:** Freiburg Institute for Musicians’ Medicine, Medical Centre University of Freiburg, University of Music Freiburg, Faculty of Medicine at the Albert-Ludwigs-University of Freiburg, Germany; Freiburg Centre for Research and Technology in Music, Germany; Tintschl BioEnergie und Strömungstechnik AG, Tintschl Unternehmensgruppe, Erlangen, Bavaria, Germany; Bamberg Symphony, Bamberg, Bavaria, Germany

**Keywords:** SARS-CoV-2 pandemic, Covid-19 virus infection, singers, dispersion of airborne transmissions, airflow, air velocity

## Abstract

Since the Covid-19 virus spreads through airborne transmission, questions concerning the risk of spreading infectious droplets during singing and music making has arisen.

To contribute to this question and to help clarify the possible risks, we analyzed 15 singing scenarios (1) qualitatively – by making airflows visible, while singing – and (2) quantitatively – by measuring air velocities at three distances (1m, 1.5m and 2m). Air movements were considered positive when lying above 0.1 m/s, which is the usual room air velocity in venue, such as the concert hall of the Bamberg Symphony, where our measurements with three professional singers (female classical style, male classic style, female popular music style) took place.

Our findings highlight that high measurements for respiratory air velocity while singing are comparable to measurements of speaking and – by far – less than coughing. All measurements for singing stayed within a reach of 1.5m, while only direct voiceless blowing achieved measurements at the 2m sensor. Singing styles that use plosive sounds, i.e. using consonants more often as in rap, produced the highest air velocities of 0.17 m/s at the 1m sensor. Also, singing while wearing a facemask produces no air movements over 0.1 m/s.

On the basis of our recent studies on measurements of airflows and air velocities of professional singers and wind instrument players, as well as further studies on CO□ measurements in room settings of music activities, we publish our results – in consideration of further up-to-date research – in our frequently updated risk assessment (first published in April 2020). On this behalf, we suggest 2m radial distances for singers, especially in choirs.

## 3. Introduction

The coronavirus pandemic has had and still has a great impact on singing, especially while performing in groups, since the SARS-CoV-2 virus infection is transported through droplets from the respiratory system and can emit from the mouth opening to the surrounding air [1]. Hence, prevailing questions concern mostly distance regulations and the fundamental questions as to whether and how individual lessons or ensemble and choir rehearsals (in different settings, levels of profession, as well as amateur choirs) can take place. Since it is known that music making in general, and therefore singing, has an extended influence on a person’s well-being [2, 3, 4], these questions concern 4 million people in Germany who sing in a choir, of which 2.2 million amateur singers are registered in a choir union [5].

In the following, we need to know at what distance air movements are being produced while singing and how these air velocities can be evaluated in comparison to usual air movements, such as speaking or coughing, as well as considering usual room air velocities.

The measurements of our study took place in the concert hall of the Bamberg Symphony (see Appendix 1) at the beginning of May 2020, when singing in choirs indoors was mainly prohibited. Up to then, there were no general recommendations on choir rehearsals, singing in smaller ensembles or single teaching situations, due to the lack of well-founded scientific medical studies.

Considering up-to-date research, there are those looking into singers and/or wind instruments, focusing either on their respiratory airflows or measuring aerosol clouds.

The study by the Ludwig-Maximilians-Universität München and the University of Erlangen [6, 7] investigated different forms of speaking and singing with 10 professional singers of the Bavarian Radio Choir. They made respiratory flows visible by exhaling the smoke of e-cigarettes (not containing nicotine). Tests were conducted in a shaded room and documented the exhaled aerosol clouds with the help of a high-speed camera and laser light. Comparing different settings (singing text, speaking text and singing without text, once with soft and once with loud phonation), they found out that singing text and speaking text reaches comparable mean distances of up to 0.85m, while singing without text reached the lowest values of 0.63m. Even though the mean measurements stayed within a reach of 1m, some singers reached distances of up to 1.4m. These measurements were then compared to coughing, which turns out to reach even further, with a mean of 1.3m and a maximum of 1.9m. On the basis of their study, they announce a suggestion for distance regulations of at least 2m to the front.

Mürbe et al. [8] pre-published a study on the increase of aerosols during professional singing. Testing 8 professional singers (two female sopranos, two female altos, two male tenors and two male baritones) of the RIAS chamber choir Berlin, they measured particle emission rates with the help of a laser particle counter during breathing, reading, singing and holding long tones. They confirmed their assumptions of singing producing higher emission rates than speaking, with mean measurements of 4.71 – 84.75 P7s during speaking and 753.4 – 6093.14 P7s during singing. Women also produced higher particle emission rates than men, leading to the assumption that high voices produce higher sound pressure level than lower voices. On the basis of their findings, they assume that singing produces more emission rates of aerosols than speaking or breathing and that an increase of aerosol rates can be observed with an increasing sound pressure level during singing, especially while holding long tones.

In consideration of their study, Mürbe et al. [9], Kriegel & Hartmann [10], and Hartmann et al. [11] published various assessments on the risk of infection with the virus-loaded aerosols, while singing indoors during the SARS-CoV-2-pandemic. They assume that different styles of singing – e.g., singing vs. speaking – as well as different intensities of voice can lead to various sizes and density of droplets and aerosols [9] and that room situations of choir rehearsals have to be taken into account [10, 11].

Furthermore, Mürbe et al. found out that child voices emit fewer aerosols during singing than adults [12]. In this study they tested 8 children (four girls and four boys) of semiprofessional children’s choirs (Staats-und Domsingknaben Berlin and a girl’s choir of the Berlin Singakademie), who were all 13 years old (except one girl was 15 years old). The study was conducted the same way as their previous study on professional adult singers as mentioned above. Their mean measurements show emission rates of 16 – 267 P/s for speaking 141 – 1240 P/s for singing, and 683 – 4332 P/s for shouting.

In 2019, before the Coronavirus infection started, the study by Asadi et al. [13] observed different speaking situations regarding their particle rate of normal speech in correlation with loudness. They support the hypotheses that coughing and sneezing are emphasizing airborne infectious disease transmissions and point out that loudness of voice is related to the particle rate of aerosols, but also assume that further factors have to be considered.

Asahi et. al furthermore emphasize that the generation of aerosols increases with the amplitude of vocalization depending on loudness, but the language spoken (English, Spanish, Mandarin, or Arabic) showed no difference.

A study conducted by the University of the German Armed Forces in Munich, which was pre-published in May 2020 [14], analyzed larger droplets when singing and speaking, as well as the flow-related small droplets when singing and playing wind instruments. The study was conducted with a professional singer, two amateur choir singers, five professional musicians (clarinet, flute, oboe, bassoon, and trumpet), and an amateur brass player (trumpet, trombone, and euphonium). This study observed the motion of droplets and air leaving either mouth or outlet during exhalation, which was then illuminated with laser and recorded with a digital camera, producing a series of images that were quantitatively analyzed afterwards. The analysis points out that while singing, airflows at a distance of 0.5m were no longer detectable regardless of volume, pitch or whether the singer was a professional or an amateur. In conclusion, they recommend a minimal radial distance of 1.5m between singers and/or wind instrumentalists.

Another recent publication [15] conducted at the Bauhaus-Universität Weimar observed the spread of exhaled air while playing wind instruments and singing using the schlieren imaging with a schlieren mirror and the background oriented schlieren method (BOS) as a way to make respiratory air visible. Two professional singers (baritone and soprano) and eleven wind instruments (woodwinds: oboe, bassoon, Bb clarinet, bass clarinet, grand flute, piccolo, alto flute; brass: Bb trumpet, tenor trombone, French horn, F tuba) of the orchestra Thüringen Philharmonic Gotha, Eisenach, were positioned in front of the schlieren mirror while playing or singing. The findings show that the spreading range, as well as the angle at which the air escapes the mouth or outlet, varies strongly from instrument and player, depending on the structure of the instrument, the structure of the mouthpiece, the way an instrument is blown and individual blowing or breathing capacities.

Based on our own studies, we updated our risk assessment for the fifth time, which was first published in April 2020 [17]. Herein we suggest a 2m radial distance between singers, as well as consideration of other factors for safe singing settings, such as social behavior and frequent ventilation to refresh CO□ emissions [18]. Our study on CO□ measurements in instrumental and vocal closed room settings showed that singers have lower emission rates than speaking or listening, leading to recommendations on frequent ventilation breaks of 5 to 10 minutes.

On behalf of this background, our study aims to provide evidence to the field of singing and music making, by providing funded data on velocity, direction and distance of respiratory air while singing different styles and pitches in comparison to speaking, as well as usual room air velocities.

## 4. Materials and Methods

### 4.1 Sample

A professional classically trained free-lance female singer of the Bamberg area, a professional female singer trained in different popular music styles of the Popakademie Baden-Württemberg, and a professional classically trained male singer of the Freiburg University of Music voluntarily took part in the study.

For the study, consent was given by the Ethics Committee of the University Medical Center Freiburg.

In order to exclude infectious persons, before the measurements the singers were questioned as to whether they had or were still suffering from typical symptoms of the Covid-19 virus disease such as a cough, loss of taste, fever or fatigue. Additionally, the temperatures of the three singers were taken with an electronic fever thermometer directly before entering the concert hall. No one reported suspicious symptoms and the measurements of the temperature showed values under the cut-off of 37,5°C in all persons. Beyond measurements, singers completely covered their mouth and nose with facemasks. The technical staff ensured mouth-nose protection by wearing facemasks during the entire measurements.

### 4.2 Design and procedure

The measurements were conducted at the beginning of May 2020 and were initiated by the Bamberg Symphony Orchestra during the first lockdown of the SARS-CoV-2-pandemic in Germany. The three professional singers were singing different styles, speaking or blowing.

For the qualitative analysis, the measurements were also filmed on a digital camera, producing videos that were qualitatively analyzed afterwards.

The different singing styles of the male classical baritone singer were an excerpt of a music piece and performing utterances including plosives. He was also tested while wearing a facemask, wearing no face mask, while speaking and while blowing directly at the sensors.

The female classical singer sang an excerpt of a classical music piece, tried out high pitches, plosive tones and blew directly in the direction of the sensors.

The female non-classically trained singer sang an excerpt of a rock song, pop lyrical, musical belting and rap. Furthermore, she was also measured while speaking and directly blowing at the sensors.

### 4.3 Flow visualization

The flow visualization by using artificial mist is one option for on-site inspection of airflows. Using the technique of flow visualization with the FlowMaker(tm), swirls have been made visible and qualitative at the mouth of the singers [19, 20].

Coming into operation was a harmless artificial fog of SAFEX®-Chemie GmbH [21] (also used for germicides in hospitals, due to containing the active component ‘Thriethylenglycol’), which consists of water droplets and is usually used as stage fog [22]. The fog droplets have a size smaller than 5µm (see Appendix 2) and can therefore be compared to the core droplets potentially carrying the SARS-CoV-2 virus [1].

The artificial fog was transported through a system called “Hydra” using a flexible tube to the release spot of the airborne particles. Through the application tube installed on a stand, the fog escaped into the free space of the room and created a cloud of fog. It was oriented towards the mouth of the singers, who sang directly into the cloud.

The movement of the fog was filmed with a video camera and qualitatively analyzed afterwards.

### 4.4 Measurement of air velocity (anemometry)

For the measurement of air velocity, omnidirectional (independent of direction) hot film probes type DISA 54N50 Low Velocity Air Flow Analyzer, manufactured by DANTEC, were used. The probes have a measuring area of 0 – 1 m/s with an accuracy of 0.2 – 0.4 % full scale. The corresponding electronics (LVFA) give a linear voltage signal of 0 – 2 Volts, according to its velocity, which is recorded with a 20 bit-AD converter on the computer. Before the measurements, the measurement chain, sensor – electronic measurement equipment – signaling cable – converter – computer, was verified in the company-owned wind tunnel using a Laser-Doppler-Anemometer (LDA).

With a distance of 1m, 1.5m and 2m from the exhaust opening, ball tubes were put on stands and placed in a line. All sensors were at a height of about 1.5m and were adjusted according to the different heights of the singers and their singing direction (see Appendices 3, 4).

The relation of the measuring signal and the actual velocity was verified according to the measurements of the in-house calibration wind tunnel in the area of 0.15 – 0.7 m/s.

### 4.5 Measurement report

Among circles of experts on air technology, room air velocities are part of indoor climate discussions. The indoor climate of habitable rooms is called comfort climate: “A climate of comfort persists when people feel thermal at ease in habitable rooms” [23]. Whether a person is feeling comfortable within a room is – amongst other components, like temperature, etc. – dependent on-air velocities, since a comfortable climate is free from draught. Draught is the undesired cooling of the body through air movement. It can be felt from a merit of 0.15 m/s and depends on the size of a room and its ventilation system [23].

The perception of comfortableness of concert halls is therefore closely related to ventilation systems of the hall. So-called “well-like ventilations” coming from the floor are usually used nowadays [24]. In floor ventilation the velocity in which the air exits the ventilation system has to be taken into account, since it is regulating how comfortable the audience feels. For concert halls, exit velocities of 0.2 m/s are recommended (in relation to a room temperature of 20°C) to not surpass an air velocity of 0.15 m/s at the height of 1m (where the audience is sitting) and to stay in the comfort zone of 0.1 – 0.2 m/s [25]. Hence, for concert halls that have a room temperature of 20°C, room air velocities of 0.1 – 0.16 m/s are usually estimated [23, 26].

Looking at the circumstances of the concert hall of the Bamberg Symphony where our measurements took place, the ventilation system was analyzed in 2017 to understand draught appearances on stage, giving our study funded data on the room air conditions, including the room temperature of 22,3°C. Hence, the draught risk – where people start to feel uncomfortable – was stated at 0.15 m/s as the limit, and therefore an area of comfortableness is considered to be from 0.1 – 0.15 m/s [25].

These numbers show that measurements under a merit of 0.1 m/s cannot be recognized by people and are comparable to “background noises”. Simultaneously, they point out that a merit of 0.15 m/s is already felt as draught, making the range of room comfortableness between 0.1 – 0.15 m/s. These numbers help to understand the measurements of the air velocities of singers, giving them a relation and comparability. Merits under 0.1 m/s are therefore not considered, while merits of 0.15 or even 0.2 m/s can be understood as remarkable.

As a basis for the analysis, charts were produced for every singer, indicating the findings of a singing scenario. These charts compare the airflow visualizations of the video sequences to the measured numbers of airflows at all the three distances (1m, 1.5m, 2m) (see Appendices 3, 4).

Making use of the descriptive analysis, the measurements of the singing scenarios were also put into a table (see Table 1), comparing air velocities of different singers and styles at the three different distances. This overview showed the maximum values of every singer at every distance, which were compared and put into a relation to the numbers of room comfortableness lying between 0.1 – 0.15 m/s. As mentioned above, a value of 0.3 m/s is felt like a strong draught, whereas a value of 0.1 m/s is not felt at all. Air velocities produced by a performer that are lying under a value of 0.1 m/s do not have an impact on the compartment air and “disappear” amongst “background noises”, meaning velocities. On the other hand, air velocities with a value of over 0.3 m/s are comparable to strong draughts or coughing and therefore have a strong impact on dispersion of air, and therefore on the dispersion of core droplets, such as SARS CoV-2 virus-carrying aerosols.

**Table 1.**
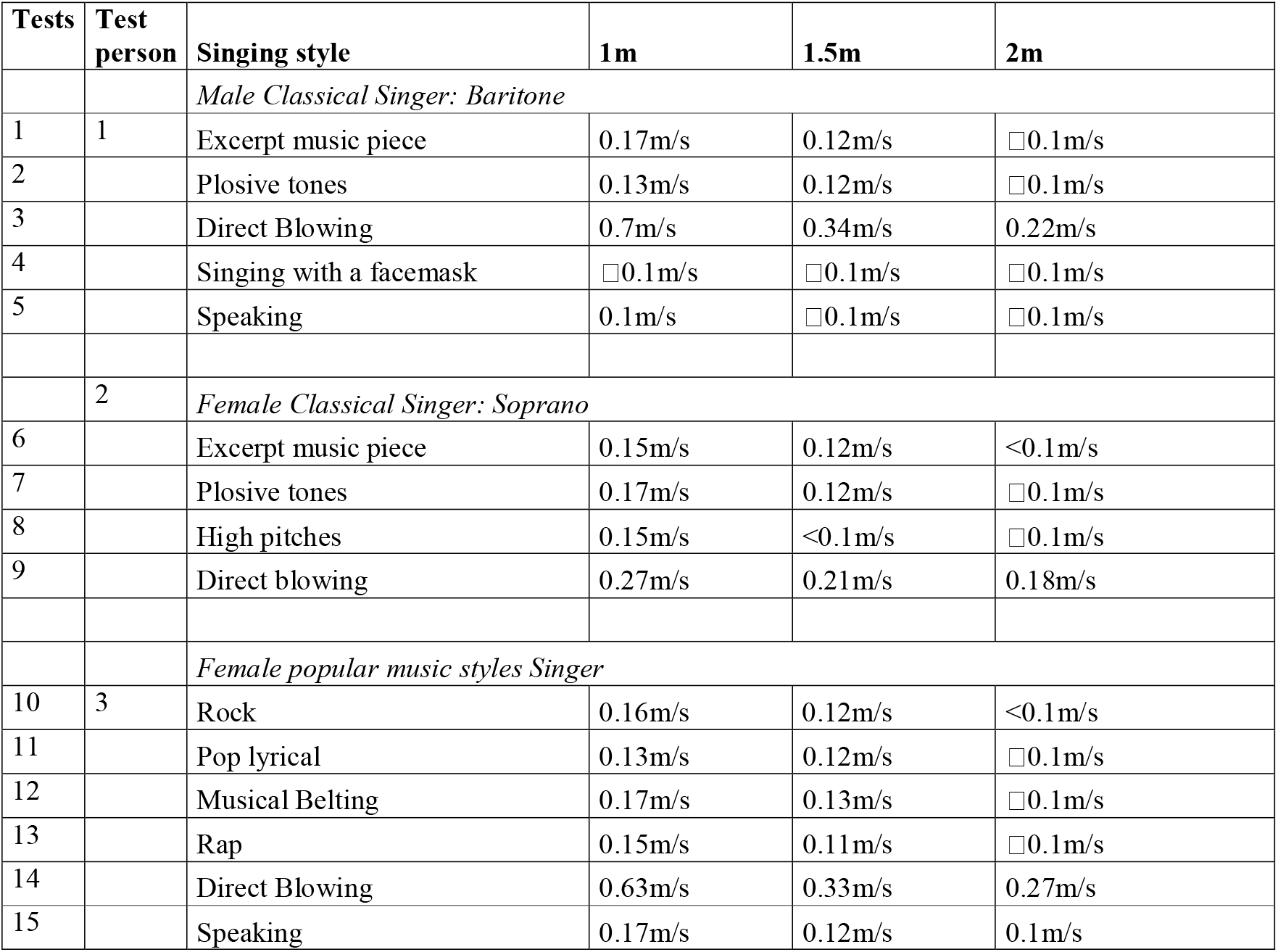
Maximum measurements of all test settings.

| Tests | Test person | Singing style | 1m | 1.5m | 2m |
| --- | --- | --- | --- | --- | --- |
|  |  | *Male Classical Singer: Baritone* |  |  |  |
| 1 | 1 | Excerpt music piece | 0.17m/s | 0.12m/s | □0.1m/s |
| 2 |  | Plosive tones | 0.13m/s | 0.12m/s | □0.1m/s |
| 3 |  | Direct Blowing | 0.7m/s | 0.34m/s | 0.22m/s |
| 4 |  | Singing with a facemask | □0.1m/s | □0.1m/s | □0.1m/s |
| 5 |  | Speaking | 0.1m/s | □0.1m/s | □0.1m/s |
|  | 2 | *Female Classical Singer: Soprano* |  |  |  |
| 6 |  | Excerpt music piece | 0.15m/s | 0.12m/s | <0.1m/s |
| 7 |  | Plosive tones | 0.17m/s | 0.12m/s | □0.1m/s |
| 8 |  | High pitches | 0.15m/s | <0.1m/s | □0.1m/s |
| 9 |  | Direct blowing | 0.27m/s | 0.21m/s | 0.18m/s |
|  |  | *Female popular music styles Singer* |  |  |  |
| 10 | 3 | Rock | 0.16m/s | 0.12m/s | <0.1m/s |
| 11 |  | Pop lyrical | 0.13m/s | 0.12m/s | □0.1m/s |
| 12 |  | Musical Belting | 0.17m/s | 0.13m/s | □0.1m/s |
| 13 |  | Rap | 0.15m/s | 0.11m/s | □0.1m/s |
| 14 |  | Direct Blowing | 0.63m/s | 0.33m/s | 0.27m/s |
| 15 |  | Speaking | 0.17m/s | 0.12m/s | 0.1m/s |

## 5. Results

### 5.1 Qualitative flow visualizations by use of artificial fog

The qualitative observations served the analysis of airflows while singing and gave a first insight into the movement of airflows while singing, speaking and blowing. Aside from that, these qualitative observations were significant to the question of whether different singing styles has an impact on the dispersion of respiratory airflows.

During the observations of singing, strong and clear air movements were seen especially while singing plosive tones or rap, and while blowing, aiming for the farthest sensor.

Only small movements were seen during singing an excerpt of a classical music piece or different modern styles.

In comparison, speaking, singing or even blowing while wearing a facemask caused no visible extraordinary airborne transmission at all. The videos of directly blowing with a face mask show that the mask lifts off the singer’s face, making visible that some air movements happen directly at the singers’ mouth, but no further movements were visible in the artificial fog.

What all qualitative observations have in common, is that they were made close to the body of the three singing persons.

Consequently, the qualitative analysis shows that singing does influence respiratory air movements, especially with regard to what is performed. Hence, the question arises on the precise measurements at different distances, and furthermore, at what distance singing can be made possible even during the SARS-CoV-2-pandemic.

### 5.2 Quantitative measurements of air velocity (anemometry)

Air velocity measurements were taken with three singers, performing a total of 15 different scenarios of singing styles and speaking (see Table 1), with and without a facemask, while singing an excerpt of a classical music piece, singing a rock song, pop lyrical, musical belting, performing a rap song, using plosive tones or directly blowing at the sensors or speaking. The numbers point out that directly blowing and aiming for one of the sensors, which is comparable to coughing, reached the highest measurements of air velocities of 0.7 m/s from the classically trained baritone singer, 0.27 m/s from the classically trained soprano singer and 0.63 m/s from the non-classically trained singer. The highest measurements for singing reached a value of 0.17 m/s while singing a classical music piece (see Appendix 5), performing high pitches or musical belting (see Appendix 6). These measurements compare to the measurements of speaking (see Table 1 and Appendices 5, 6).

#### 5.2.1 Classically trained baritone singer

##### Excerpt of music piece

When the baritone singer was singing without a facemask, maximal measurements reached values of up to 0.17 m/s at the 1m sensor. At 1.5m it reached a maximum number of 0.12 m/s, and at 2m there were no more measurements above 0.1m/s.

##### Using plosive tones

The measurements of plosive tones reached a merit of 0.13 m/s at 1m, 0.13 m/s at 1.5m and stayed below 0.1 m/s at 2m.

##### Blowing

While blowing the baritone singer did directly target the sensors at the 2m mark. Measurements reached air velocities of 0.7 m/s at 1m, 0.34 m/s at a distance of 1.5m and still had 0.22 m/s at a distance of 2m.

##### Singing and blowing with face mask

While wearing a facemask all measurements stayed under a value of 0.1m/s, independent of blowing in the direction of the sensor, singing an excerpt of a music piece, or trying out plosive tones. The measurements therefore mixed with the usual room air velocity.

##### Speaking

When the baritone singer was speaking while not wearing a facemask, measurements also stayed at 0.1 m/s, the same merit as the usual room air velocity. At a distance of 1m the measurements were reaching 0.1 m/s; at a distance of 1.5m they stayed below 0.1 m/s and showed no signal at a distance of 2m.

#### 5.2.2 Classically trained soprano singer

##### Excerpt of a music piece

While singing an excerpt of a classical music piece, the classical soprano singer sang high pitches, reaching air movement measurements of 0.15 m/s at 1m and 0.12 m/s at 1.5m, while at 2m the sensor no longer showed any air movements.

##### Plosive tones

At 1m the measurements of plosive tones reached air velocities of 0.17 m/s and at 1.5m 0.12m/s, while no air movement was measured at 2m above 0.1m/s.

##### High pitches

When the female singer tried out high pitches, air velocities at 1m showed 0.17 m/s, at 1.5m reached 0.12 m/s, and at 2m stayed under 0.1 m/s.

##### Blowing

Direct blowing to the front, aiming at the farthest mark, lead to the highest measurements of the classical soprano singer. At 1m the air velocity measurements showed up to 0.27 m/s, at 1.5m they reached 0.21 m/s, and at 2m they reached 0.18 m/s.

#### 5.2.3 Non-classically trained singer

##### Rock

While singing a part of a rock song, the measurements at 1m reached a merit of 0.16 m/s, at 1.5m they reached 0.12 m/s, and at 2m they stayed below 0.1 m/s.

##### Pop lyrical

The measurements for the pop lyrical song reached 0.13 m/s at 1m, 0.12 m/s at 1.5m, and stayed below 0.1 m/s at 2m.

##### Musical belting

When performing musical belting the measurements reached 0.17 m/s at 1m, 0.13 m/s at 1.5m and stayed below 0.1 m/s at 2m.

##### Rap

Measurements for rap reached 0.15 m/s at 1m, 0.11 m/s at 1.5m and stayed below 0.1 m/s at 2m.

##### Blowing

When the non-classical singer was directly blowing, she aimed for the farthest sensor, reaching measurements of 0.63 m/s at 1m, 0.33 m/s at 1.5m and 0.27 m/s at 2m.

##### Talking

While only talking, the non-classical singer reached merits of 0.17 m/s at a distance of 1m, 0.12 m/s at 1.5m and 0.1 m/s at 2m.

## 6. Discussion

This study observed three professional singers: a classically trained male baritone singer, a classically trained female soprano singer and a non-classically trained female singer, who all performed 15 different singing scenarios: different singing styles, speaking and blowing. Since the measurements consider various singing styles as well as comparative measurements, it has a high external validity. The findings are very representative of a high level of professional singing, with limited transferability to the amateur sector.

In order to evaluate the results of our study, in relation to recently published articles on the generation of airflows or aerosol clouds while singing, our main finding reveals that directed blowing – which is comparable to coughing – reaches much higher measurements of air velocities than singing, with the highest value of 0.7 m/s at the 1m sensor (performed by the baritone singer). At 1.5m the value dropped to 0.34 m/s and at 2m still reached a measurement of 0.22 m/s – which is a value that lies above the usual room air velocity of 0.1 – 0.15 m/s. Therefore, the risk of infection is higher while coughing rather than singing. This is a finding that is also supported by Asadi et al. [13] and Echternach et al. [6].

When comparing the measurements for singing and speaking, they show the same maximal measurements of air velocities of up to 0.17 m/s at a distance of 1m, dropping to 0.12 – 0.13 m/s at the 1.5m sensor, and staying below the usual room air velocity of 0.1m/s at the 2m sensor. In the following, for both settings of speaking and singing, air movements were no longer measurable at a distance of 2m and can therefore be considered as insignificant airflows [23].

Hence, significant airflows were measured only at 1m, with low measurements at the 1.5m sensor, which decline within or below the usual room air velocity of 0.1 – 0.15 m/s, a finding supported by Becher et al. [15], Kähler & Hain [14], Mürbe et al. [8, 12] and Echternach et al. [6].

With regard to the relation between loudness of voice and the dispersion of aerosols, our study did not find any relation, whereas other studies did, such as Asadi et al. [13] or Mürbe et al. [8, 12].

Taking into account different styles of singing, we observed that they make almost no difference at all in the generation of increasing airflows. A significant difference between blowing and singing or speaking was observed, but not by use of different singing styles. In this case, we compared mainly the highest merits of measurements of a sequence.

All qualitative observations were made close to the body of the singing person, which lead to the presumption of low measurements at the sensors, especially in distances beyond 1.5m. Quantitative measurements supported these observations, showing that respiratory air movements while singing are not dependable on volume, pitch or style of singing, but rather on individual singing habits and possibilities, which was also observed in the study of Becher et al. [15].

It is remarkable and interesting that we found differences in our assumptions after seeing the qualitative observations and having the merits from the quantitative measurements. On the basis of the qualitative observations only, it was assumed that plosive tones or rap singing would produce higher air velocities than other singing styles. The measured results showed differently: We could not find a difference if a singer is singing a song, speaking or using plosive tones – all measurements stay at the same level for all three test persons. Other studies found out otherwise, observing differences between different speaking and singing, especially while holding long tones [8], or a difference between singing with or without text, but no difference between singing or speaking text [6].

Comparing the findings on respiratory air movements while singing with our own recent study on woodwinds, it was found out that singers reach higher air velocities than woodwinds [16]. These measurements of airflows for singers stay within a reach of 1.5m, which were only surpassed during directed blowing. The highest air movement measurements for woodwinds were produced by the oboe (with a maximal value of 0.15 m/s at 1m) and the tuba (with a maximal value of 0.13 m/s at 1m), while singers reached 0.17 m/s at 1m while performing an excerpt of a music piece. Particular to instrument playing was that air-jet woodwinds, such as alto flute or piccolo, also produced side air movements of up to 0.15 m/s (alto) and 0.13 m/s (piccolo). For the other wind instruments room air movements above 0.1 m/s were no longer measurable at 1.5m while playing. They only surpassed usual room air velocities when blowing out or warming up (reaching maximal values of 0.5 m/s at 1m).

With regard to our risk assessment [17], we suggest a radial distance regulation of 2m for singers, especially when singing in groups. Only then, droplet infection due to direct contact can be excluded. This recommendation also supports the fact that strong airflows can transport heavy droplets even further than 1.5m. On the basis of our findings, though, measurements of airflows beyond 1.5m while singing were not visible. We were furthermore unable to measure jet-stream droplet effects of light droplets, which would have been visible in the artificial fog. Furthermore, we suggest frequent ventilation breaks, as mentioned in our study on CO□ measurements [18], to prevent a coronavirus infection while performing musical activities.

Aside from our findings and risk assessments, some limitations of our study have to be taken into account.

First, the study was conducted with highly professional singers, and the results can therefore not automatically be transferred to other settings or amateur singers.

Second, the test results are rather specific. We took into account different singing styles and three different singers from two different musical backgrounds. Therefore, the findings are very representative for professional classical singers as well as modern rock/pop singers, with restricted transferability to the amateur music sector and large choir groups. Since the test singers have been part of the study and know their voices very well, relevant lung capacity or ways of singing were identified for every test person. This seems to be one of the strengths of our study.

Further limitations concern the fact that the measurements were conducted with three singers only, while being sensitive to individual differences of singing, lung volume, etc. They were also performed only once for every singer, whereas more repetitions of the same sequence or analyzing more persons would have given more information on reproducibility of the test setting.

Another limitation of the measurements is the fact that air velocity measurements are very sensitive to surrounding movements, with the waving of a hand already influencing the measurements at the sensor.

## 7. Conclusion

The test results show that singing is less dangerous than coughing and comparably produces the same air velocity as speaking. In our study, results do not depend on volume, pitch or the style of singing. In this spectrum, the highest measurement of singing lies at 0.17m/s at a distance of 1m to the singer. At 2m, measurements over the usual room air velocity of 0.1m/s were no longer visible, while frontal and direct blowing, which is comparable to coughing, rise up to 0.7 m/s at the most at 1m.

Since no respiratory air movements – of any singing style – were measured at the 2m sensor, we find distance regulations of 2m to the front of the singing person to be maintainable.

In order to maintain a responsible risk assessment, we find it crucial that besides a regulated distance setup (e.g., small or large ensemble), constant fresh air ventilation and social behavior should also be taken into account. Starting from April 2020, the Freiburg Institute of Musicians’ Medicine constantly updated an official paper on “risk assessment of a coronavirus infection in the field of music” (updated May 19, July 1, July 17, and December 14, 2020), stating the findings of our various studies publicly [17] and establishing a permanent consultation on questions concerning the relationship between the coronavirus and music making.

## Data Availability

All data, mentioned in the article can be made available through the author contact.

## 8. Acknowledgements

Thanks to the singers who voluntarily took part in the study.

### [10] Appendices

**A1:**
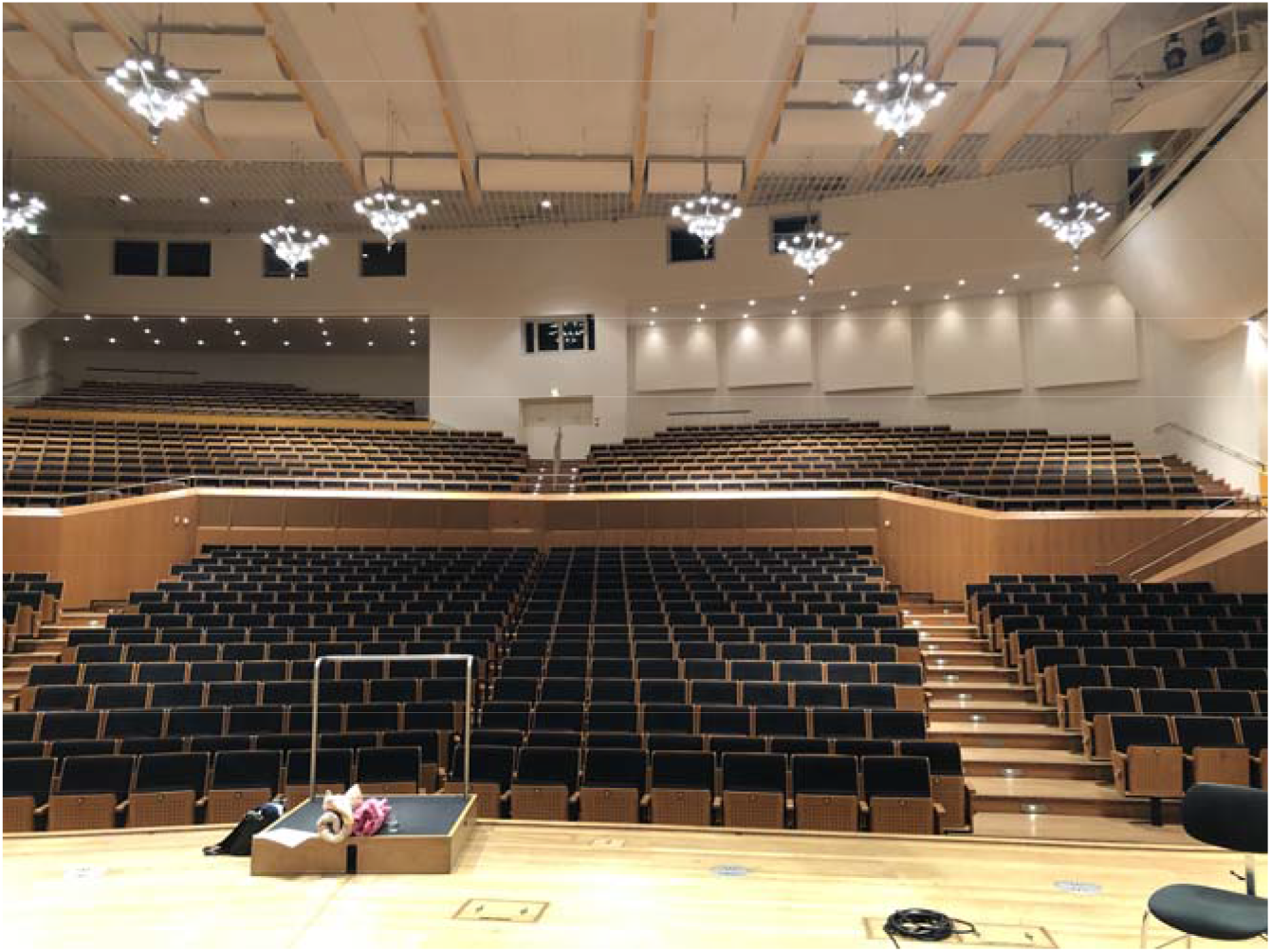
Test location: Concert hall of the Bamberg Symphony

**A2:**
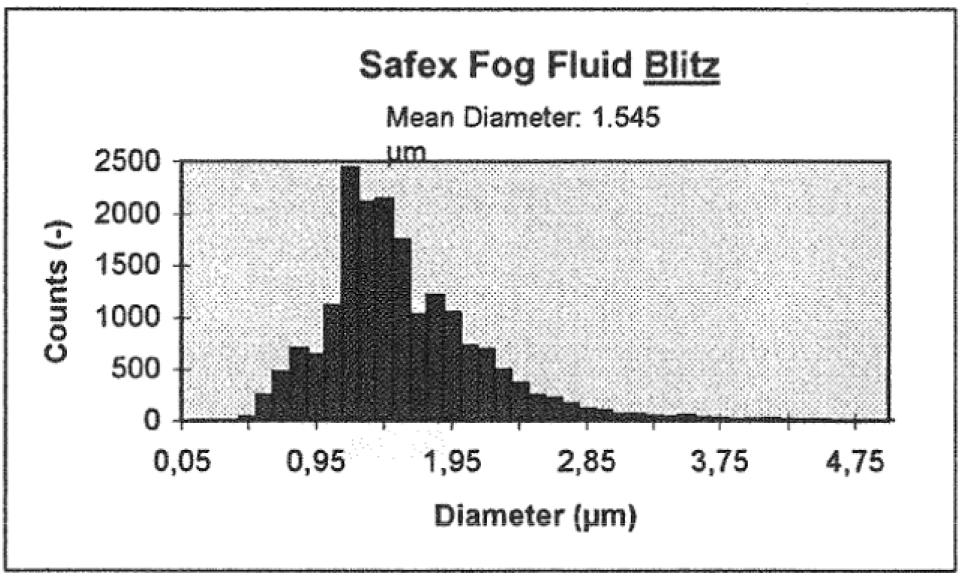
Size distribution of droplets (measurements with DANTEC PDA)

**A3:**
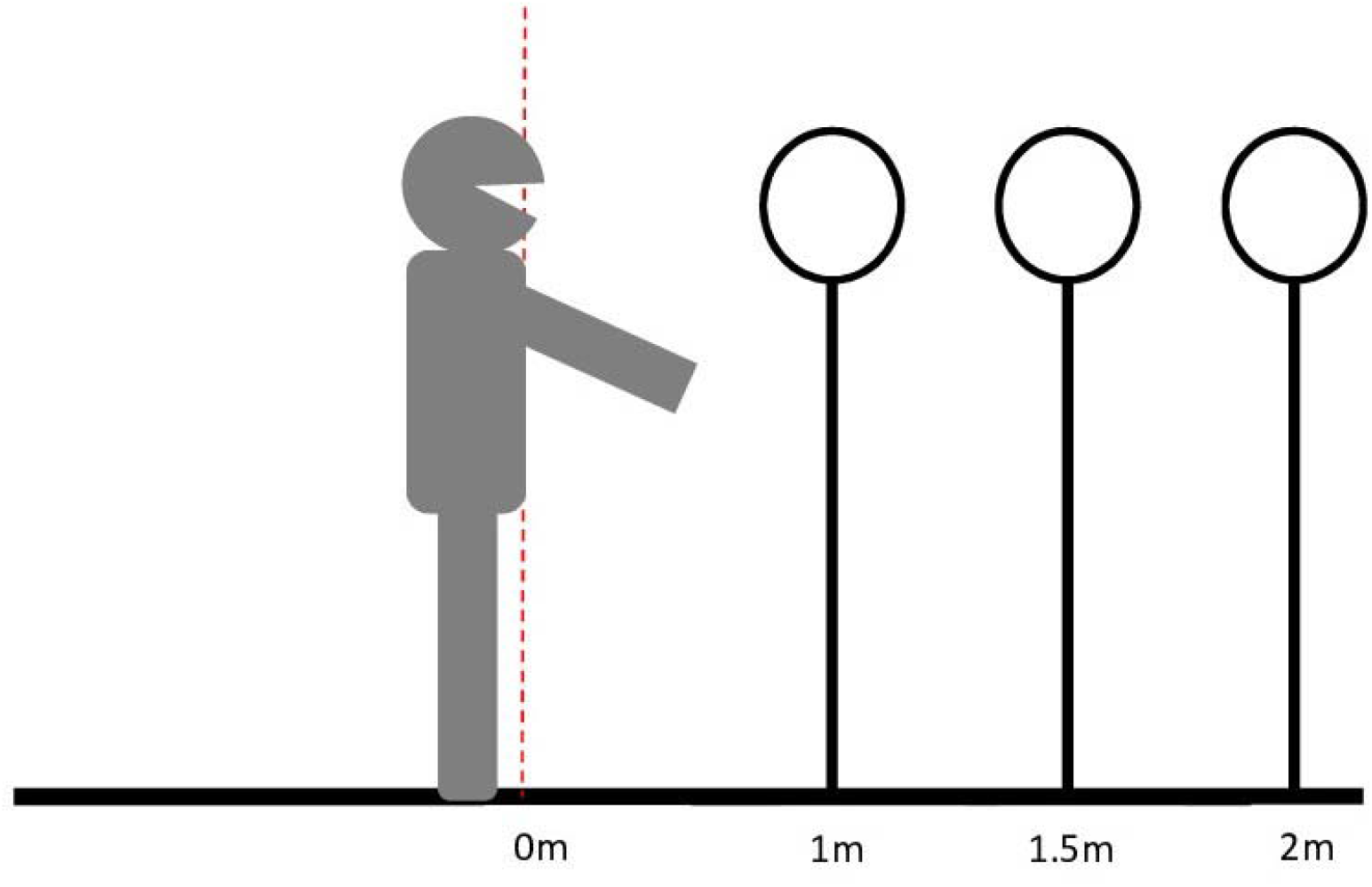
Distance measurements, while singing, at 1m, 1.5m and 2m

**A4:**
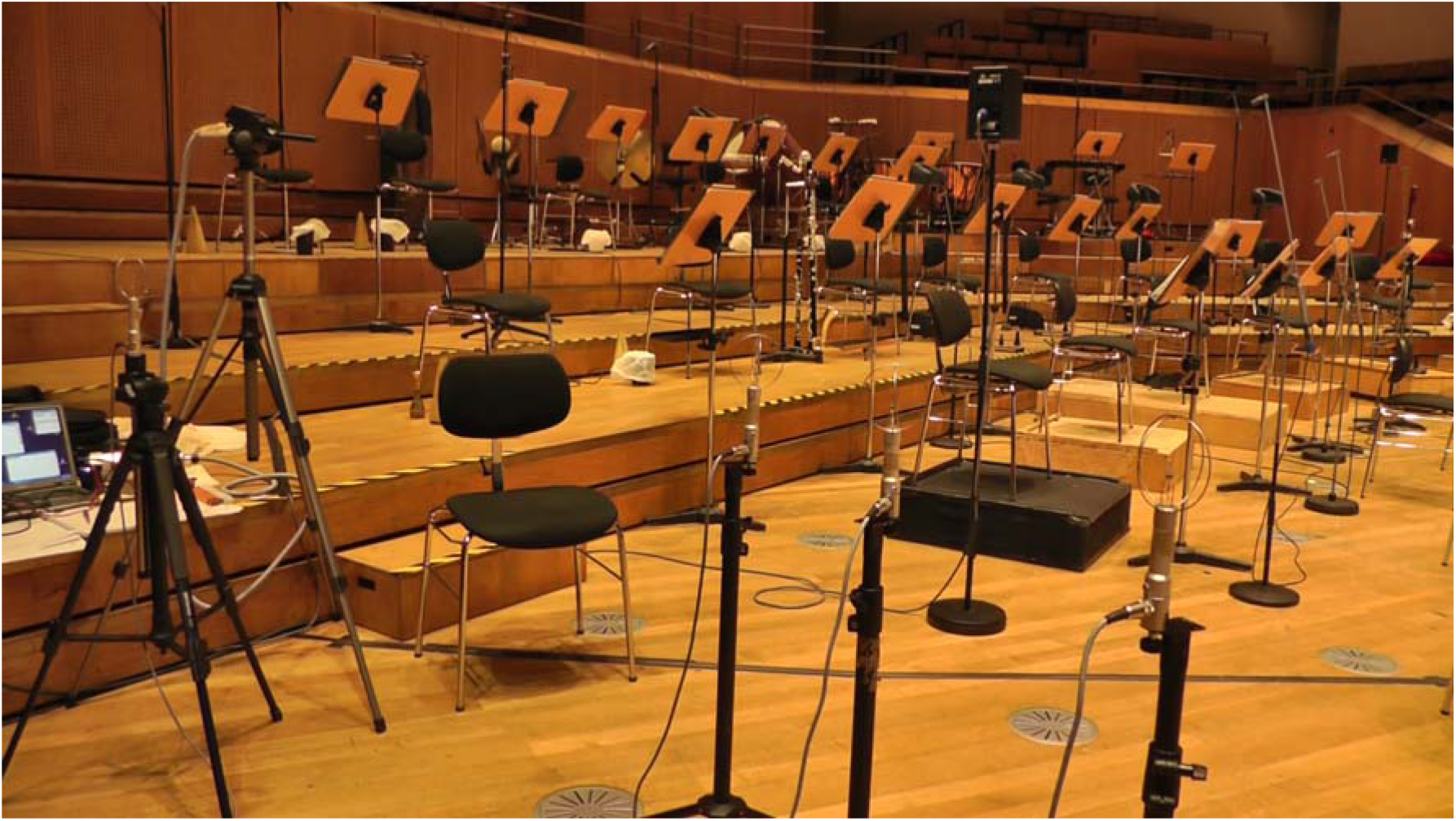
Test set-up on stage of the Bamberg Symphony hall

**A5:**
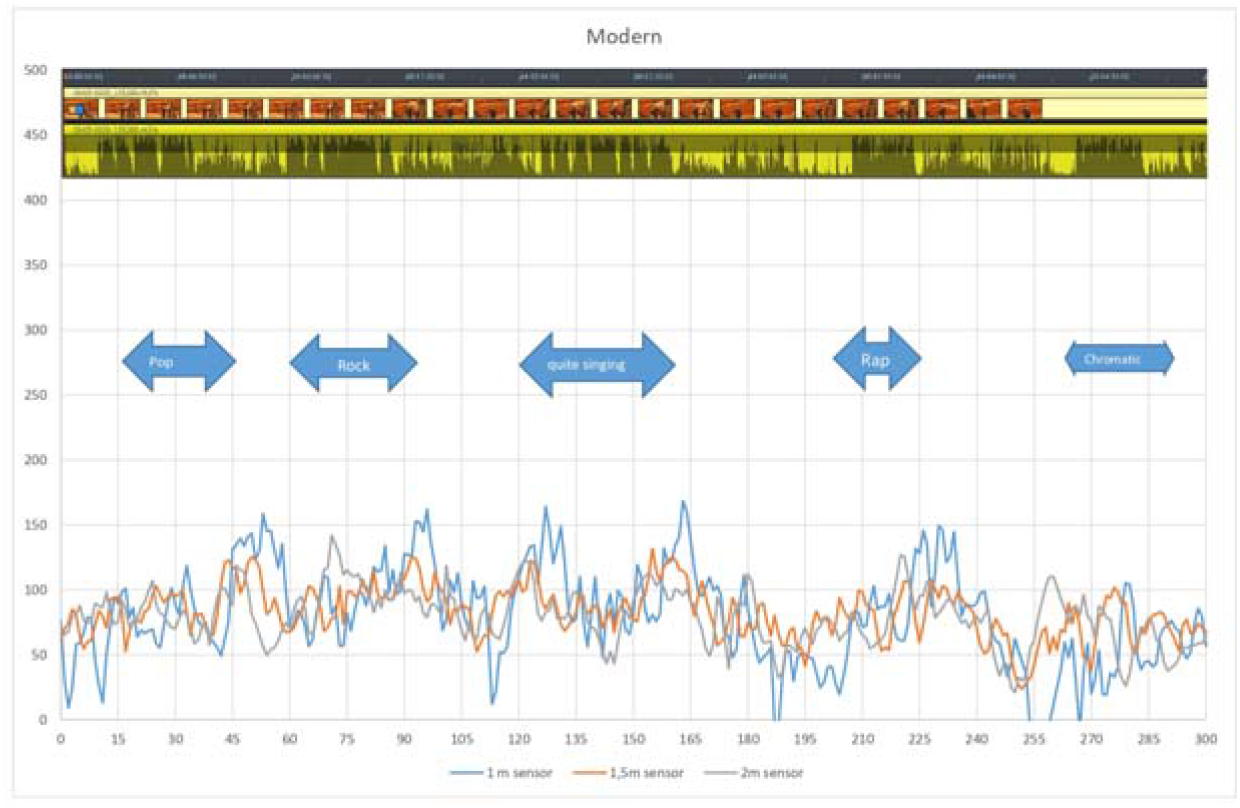
Measurements: Non-classically trained singer performing different styles

**A6:**
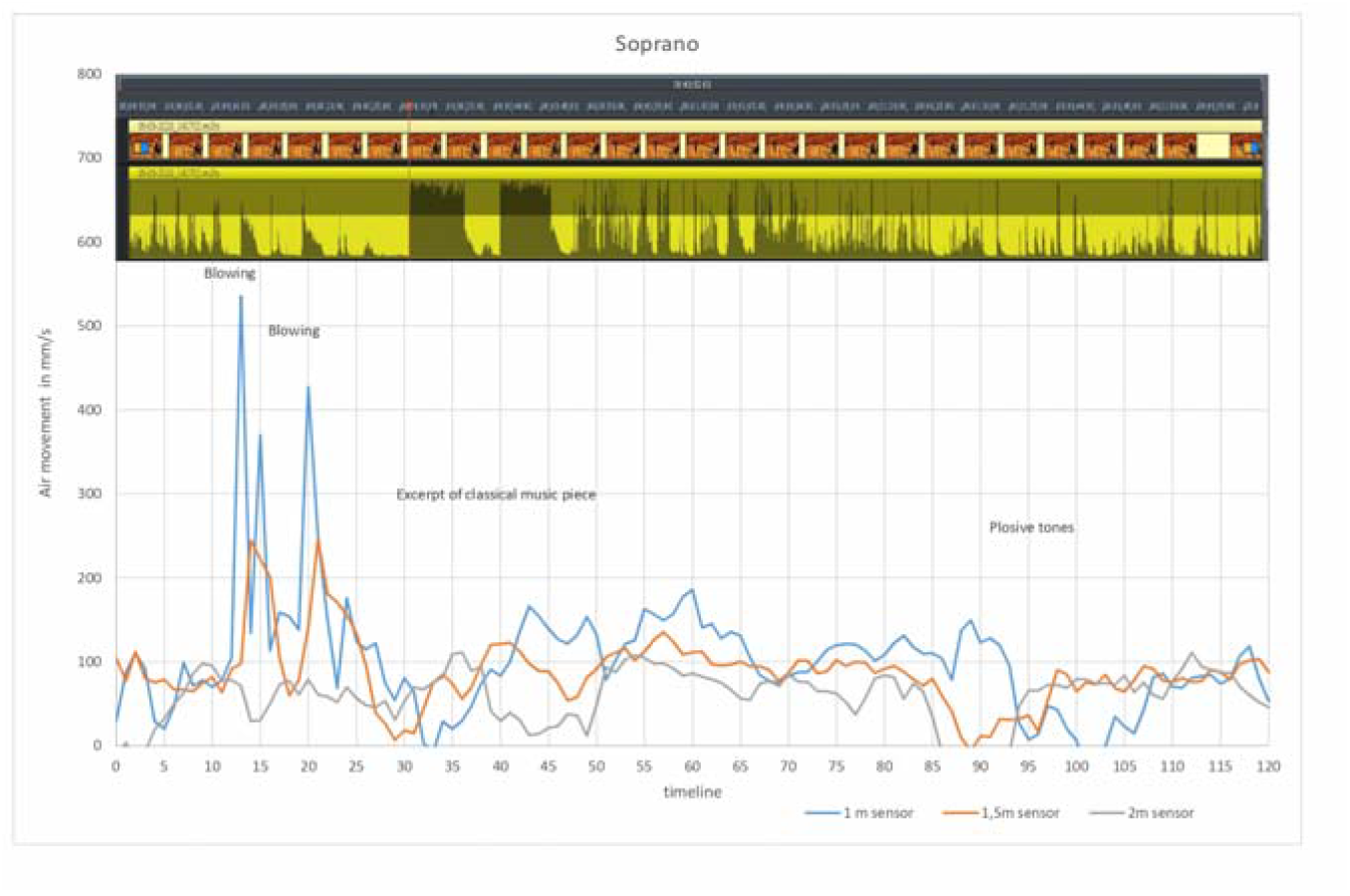
Measurements: Classically trained soprano singer

